# Factors associated with awareness and uptake of breast and cervical cancer screening among Nepalese women: Evidence from Nepal demographic and health survey 2022

**DOI:** 10.1101/2023.07.27.23293262

**Authors:** Bipul Lamichhane, Bikram Adhikari, Lisasha Poudel, Achyut Raj Pandey, Sampurna Kakchhapati, KC Saugat Pratap, Santosh Giri, Bishnu Prasad Dulal, Ishwar Tiwari, Deepak Joshi, Ghanshyam Gautam, Sushil Chandra Baral

## Abstract

**Objective:** To determine the factors associated with awareness and uptake of breast cancer screening (BCS) and cervical cancer screening (CCS) among Nepalese women aged 15-49 years.

**Methods:** We analyzed data from nationally representative Nepal demographic health survey 2022. We used weighted analysis to account for complex survey design of the survey. We presented categorical variables with frequency, percent (%) and 95% CI around percent. We used univariate and multivariable logistic regression to determine factors associated with awareness and uptake of BCS and CCS. The results of regression analysis were presented with crude odds ratio (COR) and adjusted odds ratio (AOR) and their 95% CI.

**Results:** The awareness and uptake of BCS among Nepalese women were 48.9% and 4.4% respectively whereas the awareness and uptake of CCS were 29.9% and 6.4% respectively among Nepalese women. The awareness of BCS and CCS were 1.10 and 1.22 times respectively among women with media exposure and 1.58 and 1.24 times among women with health insurance coverage. Compared to the poorest, the richest and richer have higher odds of being aware of BCS and CCS and have higher odds of BCS and CCS uptake. The uptake of CCS was 5.64 times higher among women who have heard about CCS and women who had heard about BCS had 7 times higher odds of BCS uptake.

**Conclusion:** Though awareness is relatively good, the uptake of BCS and CCS screening was very poor in Nepalese women. Provinces, ethnicity, age, education, wealth, marital status, employment, media exposure and health insurance coverage were identified as key factors associated with the awareness and uptake of BCS and CCS. These findings highlight the importance of considering socio-demographic factors in implementing effective cancer screening programs and targeting specific populations for increased awareness and uptake of screenings.

## Introduction

Globally, cancers are major public health issue and the leading cause of death, accounting for nearly 10 million deaths in 2020.^1^ In that same year, breast cancer affected 2.3 million women, tragically resulting in 685,000 deaths across the globe making it the most prevalent form of cancer. ^2^ Cervical cancer ranks as the fourth most common cancer among women on a global scale, with approximately 604,000 new cases and 342,000 deaths reported in 2020. Notably, low- and middle-income countries (LMICs) accounted for about 90% of the new cases and deaths observed worldwide during that year.^3^

The age-standardized incidence rate of cervical cancer in 2020 was estimated to be approximately 16.4 per 100,000 women, with a mortality rate of 11.1 per 100,000 women.^1^ Similarly, age-standardized incidence rate of breast cancer in 2017 was 11.54 per 100000, with a mortality rate of 6.88 per 100,000 women.^4^ Mortality rates for breast and cervical cancers continue to rise in LMICs due to late-stage diagnoses, limited access to comprehensive treatment centers, and a lack of awareness among the public.

Screening for breast and cervical cancers is a crucial strategy in preventing and detecting these types of cancer at early stages. Mammography (MMG), clinical breast examination (CBE), and breast self-examination (BSE) are the three commonly used screening tests for breast cancer. MMG is the recommended standard globally, but for low-resource settings, ^5^ CBE and BSE are also suggested due to cost-effectiveness.^6–8^ In Nepal, screening methods for cervical cancer include PAP smears, HPV testing, and visual inspection with acetic acid (VIA). Early detection of breast and cervical cancers not only saves lives but also preserves the quality of life and reduces the financial burden associated with advanced-stage treatment.^9^ However, delayed detection of breast and cervical cancers remains a common issue, particularly in low-income countries like Nepal.^10, 11^ A significant proportion of women in these countries seek medical treatment only when the cancer has reached advanced stages.^12^ A clinical study conducted in Nepal revealed that breast cancer patients often sought treatment at advanced stages, with average tumor sizes ranging from two to five centimeters. ^13^ This delay not only results in larger tumors but also complicates treatment, leading to higher mortality rates within Nepal. Additionally, the lack of universal health coverage adds a significant financial burden on affected individuals and their families.^14^

Low awareness of breast and cervical cancers and its screening among women in Nepal is a concerning issue, influenced by sociocultural dynamics and belief systems including low education levels, beliefs in witchcraft, fear of social discrimination, and stigma among others. Understanding the factors contributing to poor awareness of breast and cervical cancer in Nepal is of utmost importance. Despite frequent awareness campaigns promoting breast and cervical cancer screening, evidence suggests that women still lack awareness, necessitating further investigation into the factors that impact cancer awareness among women.^15^

To date, Nepal lacks nationally representative data on awareness and uptake of breast cancer screening (BCS) and cervical cancer screening (CCS). Therefore, this study aims to determine the status of awareness and uptake of BCS and CCS among Nepalese women, using the Nepal Demographic and Health Survey (NDHS 2022). The findings of this study will contribute to a better understanding of the awareness levels and screening practices for breast and cervical cancer in Nepal. This knowledge will then inform the development of targeted interventions and policies to improve cancer awareness and increase screening uptake._16_

## Methods

### Study design

We analyzed secondary data from NDHS 2022 in this study. NDHS 2022 is the nationally representative survey implemented by New ERA under the aegis of the Ministry of Health and Population (MoHP) with the technical support of ICF International. NDHS 2022 was funded by the US Agency for International Development (USAID).

### Study setting

Nepal is a landlocked country located in Southeast Asia with an area of 147, 516 km2. Nepal has seven administrative provinces, within which lies 753 municipalities (metropolitan cities:6, sub-metropolitan cities: 11, urban municipalities: 276, rural municipalities: 460). Nepal has three ecological belts-Mountain, Hill and Terai. Based on Census 2021, the total population of Nepal was 29164578 of which 14911027 (51.1 %) were female and 14253551 (48.9 %) were male.^17^ The human development index (HDI) of rural and urban parts of Nepal were 0.647 and 0.561 respectively and the overall HDI of Nepal was 0.587.^18^

### Sample and sampling

The sampling and sampling technique of NDHS 2022 is described elsewhere.^16^ The study used implicit stratification with proportional allocation at each lower administrative level to achieve representative sample selection. This involved sorting the sampling frame within each stratum before selecting samples based on probability-proportional-to-size at the first stage. In the first stage of sampling, 476 (248 from urban and 228 from rural) primary sampling units (PSUs) were selected with probability proportional to PSU size and with independent selection in each sampling stratum within the sample allocation. A household listing obtained from each selected PSU before the main survey served as the sampling frame for the selection of sample households in the second stage. Thirty households were selected from each cluster, for a total sample size of 14,280 households (7,440 from urban and 6,840 from rural areas). Of the total households selected, the interview was successfully accomplished in 13786 households. A total of 15238 women from 14243 households were eligible, of which 14845 women were successfully interviewed. Data generated through interviews with these 14845 women have been used for analysis in this study.

#### Data collection

Data collection for NDHS 2022 took place between January 5 and June 22, 2022. A total of 19 teams were involved in the data collection by 19 teams. and each team consisted of a supervisor, one male interviewer, three female interviewers, and one biomarker specialist.

### Dependent variables

#### Awareness of cervical and breast cancer

The woman was considered aware of cervical and breast cancer if she has heard about cervical cancer and breast cancer.

#### Awareness of cervical and breast cancer screening

The woman was considered aware of cervical and breast cancer screening if she has heard about cervical and breast cancer screening.

#### Cervical cancer screening uptake

The woman was considered to have cervical cancer screening if a doctor or other healthcare worker had ever tested her for cervical cancer.

#### Breast cancer screening uptake

The woman was considered to have breast cancer screening if a doctor or other health care worker had ever tested her for breast cancer.

### Independent variables

The independent variables assessed in this study included ecological belt (mountain, hill, Terai), setting (urban, rural), province (Koshi, Madhesh, Bagmati, Gandaki, Lumbini, Karnali, Sudurpaschim), age (in years), ethnicity (Brahmin or Chhetri, Dalit, Janajati, Madhesi, Other), religion (hindu, non-hindu), marital status (unmarried, married or living together, divorced or non-living together), wealth quintile (poorest, poorer, middle, richer, richest), education(no education, basic, secondary, higher), occupation (not working, agriculture, professional or technical or manager or clerical, sales and service, skilled or unskilled labor, others), disability (no disability, some difficulty, a lot of difficulty, can’t do at all), health insurance(covered, not covered), and media exposure (present, not present).

Women who reported reading newspapers, watching television, or listening to the radio at least once a week were categorized as having mass media exposure. Disability was assessed using the Washington Group on Disability Statistics Short Set on Functioning (WG-SS) questions^19^, which address six core functional domains: seeing, hearing, communication, cognition, walking, and self-care. These questions provide essential information on disability. If a person reported having difficulty in more than one domain, only the highest level of difficulty was considered.

### Statistical analysis

We used R version 4.2.0 ^20^ and RStudio ^21^ for data cleaning and statistical analysis. We performed a weighted analysis using “survey” package ^22^ to account complex survey design of NDHS 2022. We presented categorical variables as frequency, percent and 95% CI whereas numerical variables as mean and 95% CI. We used univariate and multivariable logistic regression to determine the association of cervical cancer screening and breast cancer screening with independent variables. The results of the logistic regression were presented as crude odds ratio and adjusted odds ratio and their 95% CI. A p-value of <0.05 was considered statistically significant.

### Ethical approval

We requested the DHS program for permission to use NDHS 2022 dataset which was granted to download and use NDHS 2022 dataset from https://www.dhsprogram.com. NHDS 2022 obtained ethical approval from the institutional review board of ICF International, United States of America (Reference number: 180657.0.001.NP.DHS.01, Date: 28^th^ April 2022) and the ethical review board of Nepal Health Research Council (Reference number: 678, Date: 30^th^ September 2021).

## Results

Table 1 illustrates the key characteristics of the Nepalese women who participated in the NDHS 2022 survey. Out of the total 14,845 women, the majority resided in urban areas (68.6%). Majority of the women were from terai region (55.1%), followed by the hilly regions (39.6%). Geographically, most women belonged to the Bagmati province (20.6%), followed by Madhesh (20.3%), Lumbini (18.1%), and Koshi (16.8%) provinces. The age range of the women spanned from 15 to 49 years, with a median age of 29 years. In terms of ethnicity, the largest group was Janajati (36.6%), followed by Brahmin/Chhetri (28.0%), Madhesi (15.7%), and Dalit (15.1%). Approximately 75.3% of the women were married, while only 4.4% had attained a higher level of education. Roughly half of the women were engaged in agriculture (48.0%), and around a quarter were unemployed (27.9%). The women were distributed equally across all wealth quintiles. About half of the women reported exposure to any form of media on a weekly basis, whereas a mere 12.0% had health insurance coverage.

**Table 1:** Characteristics of Nepalese women (N=14,845)

| Characteristic | N | % (95%CI) |
| --- | --- | --- |
| **Place of residence** |  |  |
| Urban | 10,178 | 68.6 (67.2, 69.9) |
| Rural | 4,667 | 31.4 (30.1, 32.8) |
| **Ecological region** |  |  |
| Mountain | 791 | 5.3 (3.77, 7.48) |
| Hill | 5,872 | 39.6 (35.8, 43.5) |
| Terai | 8,182 | 55.1 (51.3, 58.8) |
| **Province** |  |  |
| Koshi | 2,493 | 16.8 (15.6, 18.0) |
| Madhesh | 3,010 | 20.3 (19.1, 21.5) |
| Bagmati | 3,062 | 20.6 (19.1, 22.2) |
| Gandaki | 1,401 | 9.4 (8.33, 10.7) |
| Lumbini | 2,691 | 18.1 (17.0, 19.3) |
| Karnali | 909 | 6.1 (5.61, 6.69) |
| Sudurpaschim | 1,279 | 8.6 (7.95, 9.34) |
| **Respondent's current age, median (Q1, Q3)** | 29.0 (21.0, 38.0) |  |
| <20 years | 2,643 | 17.8 (17.1, 18.5) |
| 20-34 years | 7,216 | 48.6 (47.6, 49.6) |
| 35-49 years | 4,986 | 33.6 (32.7, 34.4) |
| **Ethnicity** |  |  |
| Brahmin/Chhetri | 4,152 | 28.0(25.8, 30.3) |
| Dalit | 2,240 | 15.1 (13.4, 16.9) |
| Janajati | 5,428 | 36.6 (33.9, 39.4) |
| Madhesi | 2,333 | 15.7 (13.6, 18.0) |
| Others | 692 | 4.7 (3.32, 6.51) |
| **Religion** |  |  |
| Hindu | 12,374 | 83.4 (81.3, 85.3) |
| Non-Hindu | 2,471 | 16.6 (14.7, 18.7) |
| **Marital status** |  |  |
| Unmarried | 3,203 | 21.6 (20.6, 22.5) |
| Married or living together | 11,180 | 75.3 (74.3, 76.3) |
| Divorced or not living together | 462 | 3.1 (2.80, 3.45) |
| **Highest educational level** |  |  |
| No education | 3,796 | 25.6 (24.1, 27.1) |
| Basic | 4,595 | 31.0 (29.7, 32.2) |
| Secondary | 5,798 | 39.1 (37.5, 40.7) |
| Higher | 656 | 4.4 (3.81, 5.11) |
| **Occupation** |  |  |
| Not working | 4,147 | 27.9 (26.2, 29.8) |
| Agriculture | 7,131 | 48.0 (45.3, 50.8) |
| Professional/technical/managerial or clerical | 1,094 | 7.4 (6.61, 8.21) |
| Sales and service | 1,222 | 8.2 (7.43, 9.12) |
| Skilled or unskilled labor | 1,235 | 8.3 (7.50, 9.22) |
| Other | 15 | 0.1 (0.04, 0.23) |
| **Wealth index** |  |  |
| Poorest | 2,628 | 17.7 (15.9, 19.7) |
| Poorer | 2,857 | 19.2 (17.6, 21.1) |
| Middle | 3,028 | 20.4 (18.9, 22.0) |
| Richer | 3,197 | 21.5 (19.9, 23.2) |
| Richest | 3,135 | 21.1 (18.7, 23.7) |
| **Covered by health insurance** | 1,775 | 12.0 (10.7, 13.4) |
| **Mass media exposure** | 7,590 | 51.1 (49.2, 53.0) |

*n*:frequency; %: percent; *CI*: confidence interval; *Q1*: First quartile, *Q3*: Third quartile

Table 2 presents the awareness of and uptake of breast and cervical cancer screening among Nepalese women. Around three fourth of the women had heard about breast cancer (75.3%) and cervical cancer (73.3%). Around half of the total women had heard about breast cancer screening whereas around one-fourth had heard about the cervical cancer screening. A total of 4.4% and 6.4% women had breast and cervical cancer screening in lifetime.

**Table 2:** Awareness of breast and cervical cancer screening, and their uptake (N = 14,845)

| Characteristic | n | % (95% CI) |
| --- | --- | --- |
| Heard of Breast cancer | 11,183 | 75.3 (73.8, 76.8) |
| Heard of Breast cancer screening | 7,259 | 48.9 (47.0, 50.8) |
| Breast cancer screening uptake | 646 | 4.4 (3.91, 4.84) |
| Heard of Cervical Cancer | 10,887 | 73.3 (71.6, 75.0) |
| Heard of Cervical Cancer screening | 4,442 | 29.9 (28.4, 31.5) |
| Cervical cancer screening uptake | 953 | 6.4 (5.73, 7.18) |

*n*:frequency; %: percent; *CI*: confidence interval

Out of those women who have heard about breast cancer, 64.9% had heard about BCS and out of those who have heard about BCS, 8.9% had BCS at least once in a lifetime. Out of those women who have heard about cervical cancer, 40.8% had heard about CCS and out of those who have heard about CCS, 21.5% had CCS screening once in a lifetime.

Table 3 presents the association of awareness of breast and cervical cancer screening with different socio-demographic characteristics. In multivariable logistic regression, the awareness of CCS was 1.52, 1.44, 1.48, 1.55, and 1.82 times higher in Bagmati, Gandaki, Lumbini, Karnali and Sudurpaschim provinces respectively compared to Koshi province. The odds of being aware of CCS was 27% and 37% lower in Madhesi, and other ethnic groups compared to the Brahmin/Chhetri. The odds of awareness of CCS was 4% higher among women aged one year older compared to women aged one year younger. The richest and richer women had 1.99 and 1.33 times higher odds of awareness of CCS compared to the poorest women. The odds of awareness of CCS were 4.49 times higher in women with higher level education compared to women without formal education. The awareness of CCS was 1.22 times among women with media exposure and 1.24 times among women with health insurance coverage. Married and divorced women had 1.54 and 1.31 times higher odds of awareness of CCS compared to unmarried women.

**Table 3:**
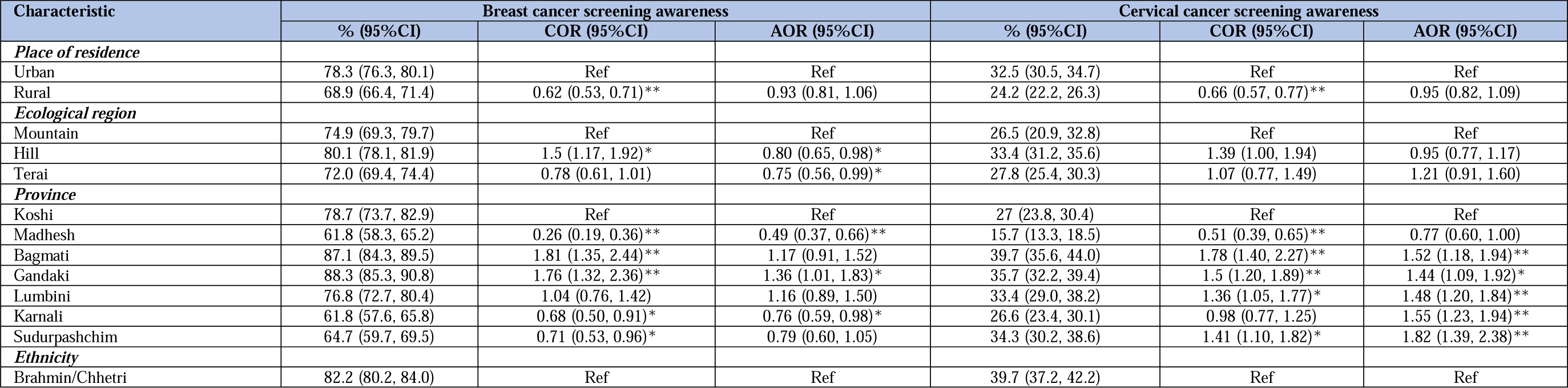
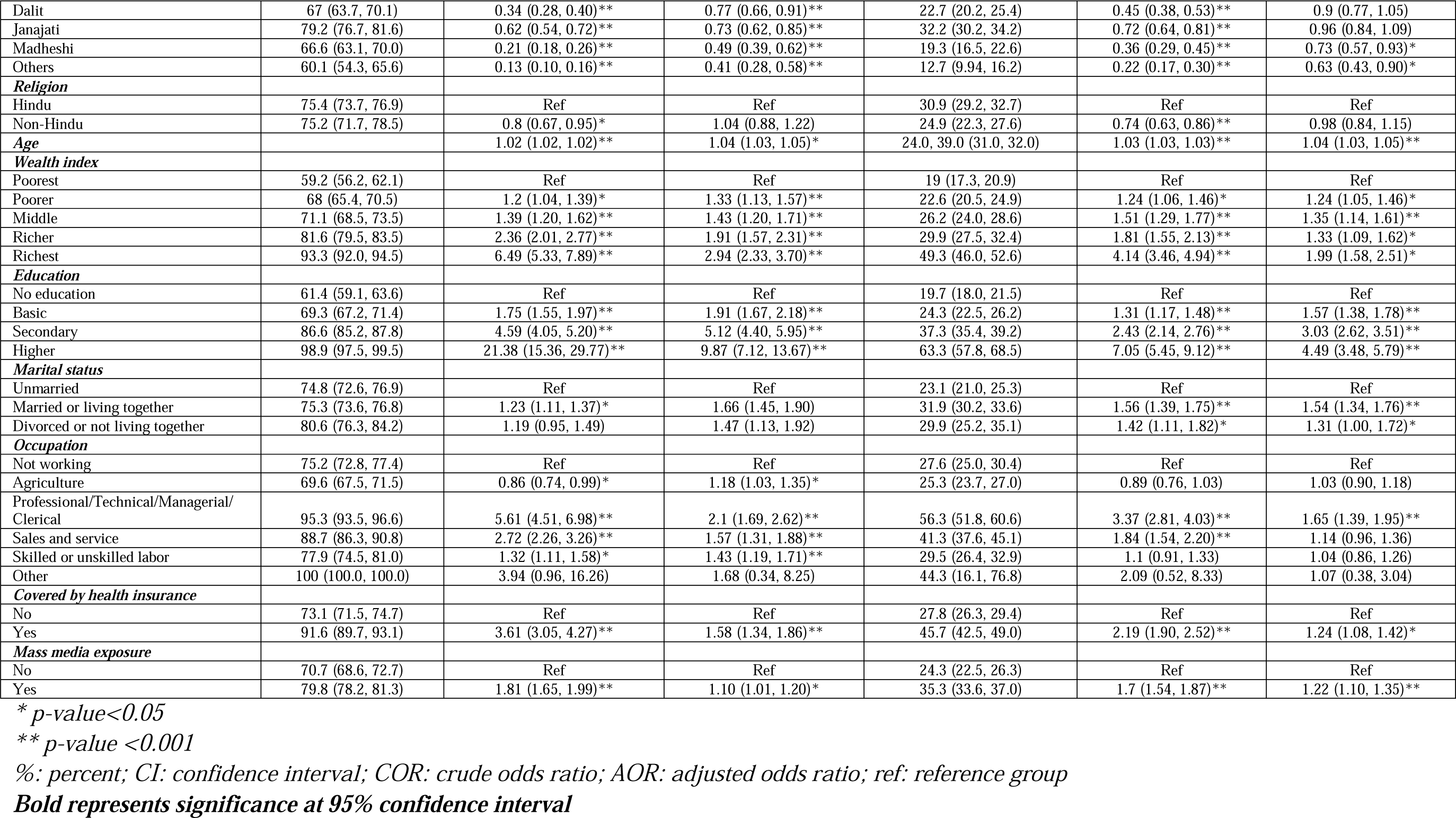
Factors associated with awareness of breast and cervical cancer screening among Nepalese women.

| Characteristic | Breast cancer screening awareness |  |  | Cervical cancer screening awareness |  |  |
| --- | --- | --- | --- | --- | --- | --- |
|  | % (95%CI) | COR (95%CI) | AOR (95%CI) | % (95%CI) | COR (95%CI) | AOR (95%CI) |
| *Place of residence* |  |  |  |  |  |  |
| Urban | 78.3 (76.3, 80.1) | Ref | Ref | 32.5 (30.5, 34.7) | Ref | Ref |
| Rural | 68.9 (66.4, 71.4) | 0.62 (0.53, 0.71)** | 0.93 (0.81, 1.06) | 24.2 (22.2, 26.3) | 0.66 (0.57, 0.77)** | 0.95 (0.82, 1.09) |
| *Ecological region* |  |  |  |  |  |  |
| Mountain | 74.9 (69.3, 79.7) | Ref | Ref | 26.5 (20.9, 32.8) | Ref | Ref |
| Hill | 80.1 (78.1, 81.9) | 1.5 (1.17, 1.92)* | 0.80 (0.65, 0.98)* | 33.4 (31.2, 35.6) | 1.39 (1.00, 1.94) | 0.95 (0.77, 1.17) |
| Terai | 72.0 (69.4, 74.4) | 0.78 (0.61, 1.01) | 0.75 (0.56, 0.99)* | 27.8 (25.4, 30.3) | 1.07 (0.77, 1.49) | 1.21 (0.91, 1.60) |
| *Province* |  |  |  |  |  |  |
| Koshi | 78.7 (73.7, 82.9) | Ref | Ref | 27 (23.8, 30.4) | Ref | Ref |
| Madhesh | 61.8 (58.3, 65.2) | 0.26 (0.19, 0.36)** | 0.49 (0.37, 0.66)** | 15.7 (13.3, 18.5) | 0.51 (0.39, 0.65)** | 0.77 (0.60, 1.00) |
| Bagmati | 87.1 (84.3, 89.5) | 1.81 (1.35, 2.44)** | 1.17 (0.91, 1.52) | 39.7 (35.6, 44.0) | 1.78 (1.40, 2.27)** | 1.52 (1.18, 1.94)** |
| Gandaki | 88.3 (85.3, 90.8) | 1.76 (1.32, 2.36)** | 1.36 (1.01, 1.83)* | 35.7 (32.2, 39.4) | 1.5 (1.20, 1.89)** | 1.44 (1.09, 1.92)* |
| Lumbini | 76.8 (72.7, 80.4) | 1.04 (0.76, 1.42) | 1.16 (0.89, 1.50) | 33.4 (29.0, 38.2) | 1.36 (1.05, 1.77)* | 1.48 (1.20, 1.84)** |
| Karnali | 61.8 (57.6, 65.8) | 0.68 (0.50, 0.91)* | 0.76 (0.59, 0.98)* | 26.6 (23.4, 30.1) | 0.98 (0.77, 1.25) | 1.55 (1.23, 1.94)** |
| Sudurpashchim | 64.7 (59.7, 69.5) | 0.71 (0.53, 0.96)* | 0.79 (0.60, 1.05) | 34.3 (30.2, 38.6) | 1.41 (1.10, 1.82)* | 1.82 (1.39, 2.38)** |
| *Ethnicity* |  |  |  |  |  |  |
| Brahmin/Chhetri | 82.2 (80.2, 84.0) | Ref | Ref | 39.7 (37.2, 42.2) | Ref | Ref |
| Dalit | 67 (63.7, 70.1) | 0.34 (0.28, 0.40)** | 0.77 (0.66, 0.91)** | 22.7 (20.2, 25.4) | 0.45 (0.38, 0.53)** | 0.9 (0.77, 1.05) |
| Janajati | 79.2 (76.7, 81.6) | 0.62 (0.54, 0.72)** | 0.73 (0.62, 0.85)** | 32.2 (30.2, 34.2) | 0.72 (0.64, 0.81)** | 0.96 (0.84, 1.09) |
| Madheshi | 66.6 (63.1, 70.0) | 0.21 (0.18, 0.26)** | 0.49 (0.39, 0.62)** | 19.3 (16.5, 22.6) | 0.36 (0.29, 0.45)** | 0.73 (0.57, 0.93)* |
| Others | 60.1 (54.3, 65.6) | 0.13 (0.10, 0.16)** | 0.41 (0.28, 0.58)** | 12.7 (9.94, 16.2) | 0.22 (0.17, 0.30)** | 0.63 (0.43, 0.90)* |
| **Religion** |  |  |  |  |  |  |
| Hindu | 75.4 (73.7, 76.9) | Ref | Ref | 30.9 (29.2, 32.7) | Ref | Ref |
| Non-Hindu | 75.2 (71.7, 78.5) | 0.8 (0.67, 0.95)* | 1.04 (0.88, 1.22) | 24.9 (22.3, 27.6) | 0.74 (0.63, 0.86)** | 0.98 (0.84, 1.15) |
| **Age** |  | 1.02 (1.02, 1.02)** | 1.04 (1.03, 1.05)* | 24.0, 39.0 (31.0, 32.0) | 1.03 (1.03, 1.03)** | 1.04 (1.03, 1.05)** |
| **Wealth index** |  |  |  |  |  |  |
| Poorest | 59.2 (56.2, 62.1) | Ref | Ref | 19 (17.3, 20.9) | Ref | Ref |
| Poorer | 68 (65.4, 70.5) | 1.2 (1.04, 1.39)* | 1.33 (1.13, 1.57)** | 22.6 (20.5, 24.9) | 1.24 (1.06, 1.46)* | 1.24 (1.05, 1.46)* |
| Middle | 71.1 (68.5, 73.5) | 1.39 (1.20, 1.62)** | 1.43 (1.20, 1.71)** | 26.2 (24.0, 28.6) | 1.51 (1.29, 1.77)** | 1.35 (1.14, 1.61)** |
| Richer | 81.6 (79.5, 83.5) | 2.36 (2.01, 2.77)** | 1.91 (1.57, 2.31)** | 29.9 (27.5, 32.4) | 1.81 (1.55, 2.13)** | 1.33 (1.09, 1.62)* |
| Richest | 93.3 (92.0, 94.5) | 6.49 (5.33, 7.89)** | 2.94 (2.33, 3.70)** | 49.3 (46.0, 52.6) | 4.14 (3.46, 4.94)** | 1.99 (1.58, 2.51)* |
| **Education** |  |  |  |  |  |  |
| No education | 61.4 (59.1, 63.6) | Ref | Ref | 19.7 (18.0, 21.5) | Ref | Ref |
| Basic | 69.3 (67.2, 71.4) | 1.75 (1.55, 1.97)** | 1.91 (1.67, 2.18)** | 24.3 (22.5, 26.2) | 1.31 (1.17, 1.48)** | 1.57 (1.38, 1.78)** |
| Secondary | 86.6 (85.2, 87.8) | 4.59 (4.05, 5.20)** | 5.12 (4.40, 5.95)** | 37.3 (35.4, 39.2) | 2.43 (2.14, 2.76)** | 3.03 (2.62, 3.51)** |
| Higher | 98.9 (97.5, 99.5) | 21.38 (15.36, 29.77)** | 9.87 (7.12, 13.67)** | 63.3 (57.8, 68.5) | 7.05 (5.45, 9.12)** | 4.49 (3.48, 5.79)** |
| **Marital status** |  |  |  |  |  |  |
| Unmarried | 74.8 (72.6, 76.9) | Ref | Ref | 23.1 (21.0, 25.3) | Ref | Ref |
| Married or living together | 75.3 (73.6, 76.8) | 1.23 (1.11, 1.37)* | 1.66 (1.45, 1.90) | 31.9 (30.2, 33.6) | 1.56 (1.39, 1.75)** | 1.54 (1.34, 1.76)** |
| Divorced or not living together | 80.6 (76.3, 84.2) | 1.19 (0.95, 1.49) | 1.47 (1.13, 1.92) | 29.9 (25.2, 35.1) | 1.42 (1.11, 1.82)* | 1.31 (1.00, 1.72)* |
| **Occupation** |  |  |  |  |  |  |
| Not working | 75.2 (72.8, 77.4) | Ref | Ref | 27.6 (25.0, 30.4) | Ref | Ref |
| Agriculture | 69.6 (67.5, 71.5) | 0.86 (0.74, 0.99)* | 1.18 (1.03, 1.35)* | 25.3 (23.7, 27.0) | 0.89 (0.76, 1.03) | 1.03 (0.90, 1.18) |
| Professional/Technical/Managerial/<br>Clerical | 95.3 (93.5, 96.6) | 5.61 (4.51, 6.98)** | 2.1 (1.69, 2.62)** | 56.3 (51.8, 60.6) | 3.37 (2.81, 4.03)** | 1.65 (1.39, 1.95)** |
| Sales and service | 88.7 (86.3, 90.8) | 2.72 (2.26, 3.26)** | 1.57 (1.31, 1.88)** | 41.3 (37.6, 45.1) | 1.84 (1.54, 2.20)** | 1.14 (0.96, 1.36) |
| Skilled or unskilled labor | 77.9 (74.5, 81.0) | 1.32 (1.11, 1.58)* | 1.43 (1.19, 1.71)** | 29.5 (26.4, 32.9) | 1.1 (0.91, 1.33) | 1.04 (0.86, 1.26) |
| Other | 100 (100.0, 100.0) | 3.94 (0.96, 16.26) | 1.68 (0.34, 8.25) | 44.3 (16.1, 76.8) | 2.09 (0.52, 8.33) | 1.07 (0.38, 3.04) |
| **Covered by health insurance** |  |  |  |  |  |  |
| No | 73.1 (71.5, 74.7) | Ref | Ref | 27.8 (26.3, 29.4) | Ref | Ref |
| Yes | 91.6 (89.7, 93.1) | 3.61 (3.05, 4.27)** | 1.58 (1.34, 1.86)** | 45.7 (42.5, 49.0) | 2.19 (1.90, 2.52)** | 1.24 (1.08, 1.42)* |
| **Mass media exposure** |  |  |  |  |  |  |
| No | 70.7 (68.6, 72.7) | Ref | Ref | 24.3 (22.5, 26.3) | Ref | Ref |
| Yes | 79.8 (78.2, 81.3) | 1.81 (1.65, 1.99)** | 1.10 (1.01, 1.20)* | 35.3 (33.6, 37.0) | 1.7 (1.54, 1.87)** | 1.22 (1.10, 1.35)** |

\* $p\text{-value} < 0.05$
\*\* $p\text{-value} < 0.001$
%; percent; CI: confidence interval; COR: crude odds ratio; AOR: adjusted odds ratio; ref: reference group

In multivariate regression analysis, the odds of being aware of BCS was 20% and 25% lower among women from hill and terai compared to mountain region. The odds of being aware of BCS among women from Madhesh and Karnali provinces were 51% and 24% lower compared to Koshi whereas 36% higher among women of Gandaki province. Compared to Brahmin, the odds of being aware of BCS was lower among all ethnic groups compared to Brahmins/Chhetri. The women of one-year older age had 4% higher odds of being aware compared to one-year younger women. The odds of being aware of BCS was 1.58 times higher in women covered with health insurance and 1.10 times higher among women with media exposure. The odds of being aware was higher among women engaged in agriculture, professional/management/clerical, sales and services, and skilled/unskilled labor compared to unemployed women. Compared to unmarried women, the odds of being aware was higher in married and divorced women. Compared to the poorest, the richest and richer had higher odds of being aware of BCS.

Table 4 presents the association between cervical and breast cancer screening uptake among Nepalese women. In the multivariable analysis, the odds of CCS uptake was 25% lower in women from rural areas compared to urban areas. The odds of CCS uptake were 1.79 times higher in women from the Madhesh province, 2.24 times higher in women from the Bagmati province, 2.13 times higher in women from the Gandaki province, and 1.65 times higher in women from the Karnali province compared to women from the Koshi province. The odds of CCS uptake were 9% higher among women who were one year older compared to women who were one year younger. The odds of CCS uptake were 1.50, 1.53, and 1.68 times higher in women with basic, primary, secondary, and higher-level education, respectively, compared to women with no formal education. The uptake was 8.64 times higher among women who had heard about CCS and 1.37 times higher among women with health insurance coverage.

**Table 4:** Factors associated with cervical and breast cancer screening uptake among Nepalese women.

| Characteristic | Breast cancer screening uptake |  |  | Cervical cancer screening uptake |  |  |
| --- | --- | --- | --- | --- | --- | --- |
|  | % (95%CI) | COR (95%CI) | AOR (95%CI) | % (95%CI) | COR (95%CI) | AOR (95%CI) |
| **Place of residence** |  |  |  |  |  |  |
| Urban | 5.0 (4.4, 5.7) | Ref | Ref | 7.7 (6.7, 8.8) |  |  |
| Rural | 2.9 (2.5, 3.4) | 0.57 (0.46, 0.71)** | 0.94 (0.73, 1.19) | 3.6 (3.1, 4.1) | 0.45 (0.36, 0.55)** | 0.75 (0.60, 0.92)* |
| **Ecological region** |  |  |  |  |  |  |
| Mountain | 4.2 (2.6, 6.6) | Ref | Ref | 5.5 (3.9, 7.7) | Ref | Ref |
| Hill | 5.7 (4.9, 6.6) | 1.39 (0.83, 2.35) | 0.85 (0.61, 1.17) | 8.5 (7.2, 9.9) | 1.58 (1.06, 2.37) | 1.13 (0.70, 1.82) |
| Terai | 3.4 (2.9, 4.0) | 0.81 (0.48, 1.36) | 0.63 (0.43, 0.94) * | 5 (4.1, 6.1) | 0.91 (0.60, 1.37) | 1.12 (0.66, 1.92) |
| **Province** |  |  |  |  |  |  |
| Koshi | 3.7 (2.8, 5.0) | Ref | Ref | 4.2 (3.4, 5.1) | Ref | Ref |
| Madhesh | 2.6 (1.8, 3.7) | 0.68 (0.42, 1.10) | 1.34 (0.79, 2.29) | 3.4 (2.4, 5.0) | 0.82 (0.53, 1.27) | 1.79 (1.10, 2.94)* |
| Bagmati | 7.3 (5.9, 9.0) | 2.01 (1.37, 2.96)** | 1.2 (0.79, 1.81) | 12.1 (9.6, 15.1) | 3.17 (2.27, 4.41)** | 2.24 (1.51, 3.32)** |
| Gandaki | 6.4 (5.3, 7.8) | 1.76 (1.22, 2.56)* | 1.16 (0.76, 1.76) | 9.6 (8.1, 11.4) | 2.44 (1.84, 3.25)** | 2.13 (1.47, 3.07)** |
| Lumbini | 3.8 (2.9, 4.9) | 1.01 (0.67, 1.52) | 1.04 (0.70, 1.55) | 5.2 (4.0, 6.8) | 1.26 (0.88, 1.79) | 1.24 (0.90, 1.71) |
| Karnali | 3 (2.3, 3.9) | 0.79 (0.52, 1.20) | 1.05 (0.66, 1.66) | 4.9 (3.5, 6.7) | 1.18 (0.79, 1.76) | 1.65 (1.02, 2.67)* |
| Sudurpashchim | 2.7 (1.9, 3.6) | 0.7 (0.45, 1.09) | 1.01 (0.65, 1.58) | 4.4 (3.4, 5.7) | 1.07 (0.76, 1.50) | 1.14 (0.77, 1.67) |
| **Ethnicity** |  |  |  |  |  |  |
| Brahmin or chhetri | 5.8 (4.9, 6.8) | Ref | Ref | 10.2 (8.7, 11.8) | Ref | Ref |
| Dalit | 3.6 (2.8, 4.6) | 0.6 (0.45, 0.81)** | 1.55 (1.10, 2.17)* | 4 (3.1, 5.1) | 0.37 (0.27, 0.49)** | 0.83 (0.59, 1.17) |
| Janajati | 4.5 (3.7, 5.4) | 0.76 (0.58, 0.99) | 1.12 (0.83, 1.49) | 6.4 (5.5, 7.4) | 0.60 (0.50, 0.73)** | 0.81 (0.63, 1.04) |
| Madheshi | 3.0 (2.1, 4.2) | 0.5 (0.34, 0.75) | 1.35 (0.88, 2.08) | 3.6 (2.7, 4.8) | 0.33 (0.23, 0.46)** | 0.68 (0.45, 1.04) |
| Others | 2.0 (1.1, 3.8) | 0.34 (0.17, 0.66) | 1.57 (0.69, 3.56) | 1.7 (0.8, 3.6) | 0.15 (0.07, 0.34)** | 0.46 (0.19, 1.13) |
| **Religion** |  |  |  |  |  |  |
| Hindu | 4.5 (4.0, 5.0) | Ref | Ref | 6.7 (5.9, 7.5) | Ref | Ref |
| Non-hindu | 3.8 (3.0, 4.8) | 0.84 (0.65, 1.09) | 0.97 (0.73, 1.29) | 5.2 (4.1, 6.6) | 0.77 (0.60, 0.99) | 1.09 (0.80, 1.49) |
| **Age** | 42.0 (34.0, 37.0) | 1.06 (1.05, 1.07)** | 1.05 (1.04, 1.06)** | 43.0 (38.0, 39.0) | 1.1 (1.09, 1.11)** | 1.09 (1.07, 1.10)** |
| **Wealth index** |  |  |  |  |  |  |
| Poorest | 1.9 (1.4, 2.4) | Ref | Ref | 3.1 (2.6, 3.8) | Ref | Ref |
| Poorer | 3.1 (2.40, 3.88) | 1.66 (1.16, 2.36) | 1.4 (0.97, 2.01) | 3.7 (3.0, 4.6) | 1.2 (0.88, 1.64) | 0.93 (0.66, 1.32) |
| Middle | 3.1 (2.49, 3.93) | 1.7 (1.19, 2.43) | 1.3 (0.88, 1.92) | 4.1 (3.4, 5.0) | 1.33 (1.01, 1.75) | 0.87 (0.61, 1.24) |
| Richer | 3.9 (3.25, 4.79) | **2.16 (1.55, 3.02)\*\*** | **1.24 (0.83, 1.84)** | **5.4 (4.6, 6.3)** | **1.75 (1.36, 2.26)\*\*** | **0.91 (0.63, 1.31)** |
| Richest | 9.2 (7.84, 10.8) | **5.33 (3.87, 7.35)\*\*** | **1.63 (1.06, 2.51)\*** | **14.9 (12.5, 17.7)** | **5.45 (4.09, 7.28)\*\*** | **1.26 (0.82, 1.92)** |
| **Education** |  |  |  |  |  |  |
| No education | 2.8 (2.21, 3.43) | **Ref** | **Ref** | **5 (4.2, 5.9)** | **Ref** | **Ref** |
| Basic | 3.8 (3.16, 4.50) | **1.39 (1.04, 1.84)\*** | **1.37 (1.00, 1.88)\*** | **5.8 (5.0, 6.8)** | **1.18 (0.93, 1.50)** | **1.50 (1.13, 1.98)\*** |
| Secondary | 4.9 (4.26, 5.66) | **1.82 (1.41, 2.35)\*\*** | **1.55 (1.11, 2.17)\*** | **6.5 (5.4, 7.8)** | **1.32 (1.02, 1.72)\*** | **1.53 (1.13, 2.08)\*** |
| Higher | 12.6 (9.52, 16.6) | **5.1 (3.45, 7.55)\*\*** | **2.00 (1.22, 3.27)\*** | **18 (14.1, 22.6)** | **4.17 (3.00, 5.78)\*\*** | **1.68 (1.00, 2.80)\*** |
| **Marital status** |  |  |  |  |  |  |
| Unmarried | 1.1 (0.7, 1.7) | **Ref** | **Ref** | **0.2 (0.1, 0.6)** | **Ref** | **Ref** |
| Married or living together | 5.2 (4.7, 5.9) | **4.96 (3.08, 7.97)\*\*** | **3.20 (1.87, 5.47)** | **8.2 (7.3, 9.3)** | **49.15 (15.29, 158.04)\*\*** | **19.63 (5.99, 64.30)\*\*** |
| Divorced or not living together | 5.6 (3.6, 8.6) | **5.32 (2.78, 10.18)\*\*** | **3.35 (1.60, 7.03)** | **5.7 (3.7, 8.8)** | **33.25 (9.31, 118.69)\*\*** | **9.67 (2.49, 37.52)\*\*** |
| **Occupation** |  |  |  |  |  |  |
| Not working | 4.1 (3.3, 5.0) | **Ref** | **Ref** | **5.5 (4.4, 6.8)** | **Ref** | **Ref** |
| Agriculture | 3.0 (2.61, 3.54) | **0.73 (0.57, 0.95)\*** | **0.75 (0.57, 0.99)\*** | **4.8 (4.2, 5.4)** | **0.87 (0.67, 1.13)** | **0.93 (0.71, 1.21)** |
| Professional, manager, clerical | 10.5 (8.43, 13.0) | **2.75 (2.02, 3.74)\*\*** | **1.19 (0.88, 1.61)** | **15 (12.0, 18.6)** | **3.04 (2.37, 3.90)\*\*** | **1.23 (0.90, 1.69)** |
| Sales and service | 7.3 (5.6, 9.4) | **1.83 (1.29, 2.60)\*\*** | **0.96 (0.68, 1.36)** | **12.1 (9.8, 14.8)** | **2.36 (1.78, 3.15)\*\*** | **1.21 (0.90, 1.62)** |
| Skilled/unskilled labor | 4.4 (3.1, 6.2) | **1.08 (0.70, 1.68)** | **0.84 (0.55, 1.29)** | **5.5 (4.2, 7.3)** | **1.01 (0.71, 1.43)** | **0.84 (0.58, 1.24)** |
| Other | 7.1 (1.4, 29.1) | **1.78 (0.32, 9.94)** | **0.58 (0.09, 3.72)** | **25.3 (3.6, 75.3)** | **5.82 (0.67, 50.47)** | **1.15 (0.21, 6.34)** |
| **Covered by health insurance** |  |  |  |  |  |  |
| No | 3.8 (3.4, 4.3) | **Ref** | **Ref** | **5.6 (4.9, 6.3)** | **Ref** | **Ref** |
| Yes | 8.4 (6.9, 10.3) | **2.34 (1.82, 2.99)\*** | **1.28 (0.99, 1.65)** | **12.7 (10.7, 14.9)** | **2.46 (2.01, 3.01)\*\*** | **1.37 (1.09, 1.71)\*** |
| **Mass media exposure** |  |  |  |  |  |  |
| No | 2.9 (2.5, 3.5) | **Ref** | **Ref** | **4.6 (3.9, 5.4)** | **Ref** | **Ref** |
| Yes | 5.7 (5.0, 6.4) | **1.99 (1.64, 2.41)** | **1.39 (1.13, 1.71)** | **8.1 (7.2, 9.2)** | **1.84 (1.53, 2.21)** | **1.18 (0.95, 1.46)** |
| **Heard about BCS or CCS** |  |  |  |  |  |  |
| No | 0.8 (0.6, 1.1) | **Ref** | **Ref** | **3.0 (2.6, 3.6)** | **Ref** | **Ref** |
| Yes | 8.1 (7.3, 8.9) | **10.84 (7.84, 14.99)\*\*** | **7.29 (5.38, 9.88)\*\*** | **17.5 (15.7, 19.5)** | **12.5 (10.24, 15.25)\*\*** | **8.15 (6.62, 10.03)\*\*** |

\* $p\text{-value} < 0.05$
\*\* $p\text{-value} < 0.001$
%; percent; CI: confidence interval; COR: crude odds ratio; AOR: adjusted odds ratio; ref: reference group

In the multivariable logistic regression to determine the association of BCS uptake with different socio-demographic variables, the odds of BCS uptake were 37% lower among women from the Terai compared to women from the mountain region. The odds of BCS uptake were 1.55 times higher in Dalit women compared to Brahmin/Chhetri women. The odds of BCS uptake were 5% higher among women who were one year older compared to women who were one year younger. Compared to the poorest women, the richest women had 1.63 times higher odds of BCS uptake. The odds of BCS uptake were 1.27, 1.55, and 2.00 times higher in women with basic, primary, secondary, and higher-level education, respectively, compared to women with no formal education. Married and divorced women had higher odds of BCS uptake compared to unmarried women. Women who had heard about BCS had 7 times higher odds of BCS uptake. The odds of uptake of BCS among women engaged in agriculture was 25% lower compared to unemployed women.

## Discussion

This study assessed the factors associated with awareness of breast and cervical cancer screening and uptake among Nepalese women using secondary data from NDHS 2022. Around three fourth of the women had heard about breast cancer (75%) and cervical cancer (73%). Around half of the total women had heard about breast cancer screening whereas around one-fourth had heard about cervical cancer screening.

The multivariate analysis revealed that breast and cervical cancer screening awareness exhibited positive associations with certain factors. These factors included belonging to the Brahmin/Chhetri ethnic group, older age group, having a higher socioeconomic status (specifically, being from richer households), possessing a higher level of education, having exposure to media, being covered by health insurance, and being married. The study also examined the association between cervical and breast cancer screening uptake among Nepalese women. For cervical cancer screening (CCS) uptake, women from urban areas had higher uptake rates compared to rural areas. Women from certain provinces, such as Madhesh, Bagmati, Gandaki, and Karnali, had higher odds of CCS uptake than women from Koshi province. CCS uptake increased with age, education level, awareness of CCS, and health insurance coverage. Regarding breast cancer screening (BCS) uptake, women from the mountain region had higher uptake rates than women from the Terai region. Dalit women showed higher odds of BCS uptake compared to Brahmin/Chhetri women. BCS uptake increased with age, education level, awareness of BCS, and marital status (married and divorced women had higher odds compared to unmarried women). However, women engaged in agriculture had lower odds of BCS uptake compared to unemployed women.

The awareness level of breast cancer reported in the current study (75.3%) falls within the range observed in various regions worldwide. Studies conducted in different countries have reported a wide range of breast cancer awareness levels, typically varying from around 40% to 90%.^23–29^ Awareness levels tend to be higher in more developed countries and regions with robust healthcare systems and greater access to information and health services. The findings from two studies conducted in Nepal demonstrated that a substantial proportion of respondents had heard about cervical cancer, with awareness levels reported at 88.8% and 77.5%, respectively Likewise, in Nepal, Nigeria, Uganda, and India, several other studies reveal that the percentage of women who possess knowledge about cervical cancer ranged from 65% to 99%. ^30–33^ The relatively low awareness of breast cancer among women identified in our study underscores the urgent need for targeted awareness promotion, particularly among vulnerable and at-risk populations. Improving awareness of breast cancer is crucial in fostering better screening habits and early detection, which are fundamental to enhancing the effectiveness of cancer treatment.

Consistent with prior research, our study also demonstrates a positive association between the increasing age of women and their awareness of breast cancer (BC) ^34,35^ and cervical cancer (CC),^36^ as well as their awareness of breast cancer screening (BCS)^37^ and cervical cancer screening (CCS).^38^ Traditionally, education initiatives focused on BC and CC have predominantly targeted populations deemed to be at higher risk of developing these cancers. However, recent evidence highlights the importance of extending screening efforts to include younger women as well.^39^ Despite breast cancer screening being more prevalent among women in the premenopausal and menopausal age groups, it is crucial to recognize the increasing incidence of these diseases among younger populations. Therefore, it is essential to direct health education and BC and CC awareness promotion towards this demographic. This consideration is particularly critical in low- and middle-income countries (LMICs) where healthcare infrastructure may not be fully developed to handle advanced stages of these diseases. Consequently, early BC and CC education assume paramount importance, especially among women in their reproductive years. By emphasizing early detection and treatment, women can be better equipped to address the challenges posed by BC and CC, leading to improved health outcomes.^39, 40^

The findings of our study align with previous investigations conducted in Ethiopia,^41^ Uganda,^42^ and Lesotho^35^ which have consistently shown that individuals who had access to television or radio were more likely to be aware of breast cancer screening (BCS) and cervical cancer screening (CCS). This highlights the critical role of media information in improving awareness levels related to breast cancer. To maximize the reach and impact of health education efforts, it is crucial to utilize diverse and multiple channels to disseminate information about breast cancer. Both print and electronic media have proven to be effective tools in health education initiatives. ^35^

The present study’s findings concerning breast cancer (BC) and cervical cancer (CC) screening uptakes are consistent with prior research, corroborating the importance of knowledge acquisition in influencing screening utilization. The estimated uptake of cervical cancer screening (CCS) among women in Ethiopia, as reported by Kassie et al., was found to be 8%. Moreover, their study highlights the significant association between knowledge of cervical cancer and the utilization of CCS. Mahumud et al. examined the uptake of breast cancer screening (BCS) in low-resource Asian countries and reported an overall utilization rate of 19%. Similar to our study, Mahumud et al. identified several factors associated with increased BCS uptake, including higher education levels, older age at first birth, female-headed households, access to media communication, and urban residency. Similarly, Islam et al. conducted a systematic review, demonstrating that screening practices for both breast and cervical cancers were positively correlated with opportunities for knowledge acquisition. Similar to our study, Islam, et al. also showed that women with higher education levels, urban residency, and employment outside the home exhibited increased screening behaviors. These determinants reflect the complex interplay of sociocultural and economic factors that shape health-seeking behaviors, especially in resource-constrained regions. The findings highlight the importance of designing multifaceted interventions that address both knowledge gaps and socioeconomic barriers to improve breast cancer and cervical cancer screening rates.

### Strengths and limitations

This study has several strengths. First, this study is based on the nationally representative survey data so, the results of the study can be generalized over Nepal. Second, this study employed validated questionnaire and trained data enumerators to collect data. Third, we apply weighted analysis to account non-response and complex survey design. Some limitations linked to this study included a) the directionality of the association of awareness and uptake of screening can’t be established due to cross-sectional nature of this study, b) awareness and uptake of cervical and breast cancer screening was self-reported which could be affected by recall-bias and social desirability.

### Implication to policy makers

In the context of Nepal, enhancing breast cancer screening awareness, particularly among vulnerable and at-risk populations, is pivotal to improving early detection and screening practices. Policymakers should prioritize health promotion interventions that target rural communities, women without formal education, women from poorest wealth quintile to raise awareness and uptake of breast and cervical cancer screening. The findings from this study are valuable for clinicians and policymakers engaged in developing strategies for promoting breast and cervical cancer screening awareness and their uptake.

## Conclusion

Though awareness of breast and cervical cancer is higher, the awareness and uptake of BCS and CCS were relatively lower among Nepalese women. Awareness and uptake of BCS and CCS are associated with mass media exposure, enrollment in health insurance program, age of the women, marital status, education attainment and wealth quintile. The findings emphasize the need for targeted awareness campaigns, continued efforts in health promotion and education, and comprehensive interventions to address these socio-demographic disparities and improve the awareness and uptake of breast and cervical cancer screenings. By tailoring strategies to address the identified factors, healthcare authorities can effectively increase awareness and utilization of screenings, leading to early detection and improved management of breast and cervical cancer cases.

## Data Availability

The data can be downloaded after receiving approval from the DHS program (https://www.dhsprogram.com).

